# Temporal Trends in Lifestyle Modification of Patients with Hypertension in Korea, 1998–2021

**DOI:** 10.1101/2023.08.22.23294451

**Authors:** Jun Hwan Cho, Gyu Tae Park, Kyung-Taek Park, Hyue Mee Kim, Sungsoo Cho, Sang Yeub Lee, Young-Hoon Jeong, Wang-Soo Lee, Sang-Wook Kim, Hoyoun Won

## Abstract

**Background:** Lifestyle modification is crucial in managing hypertension, independent of medical treatment. This study aimed to evaluate the implementation of lifestyle modifications and analyze the trends in lifestyle modification among patients with hypertension in Korea over the past two decades.

**Methods:** We analyzed data from the Korea National Health and Nutrition Examination Survey (KNHANES) conducted between 1998 and 2021. The study included adults aged ≥ 20 years. Factors such as regular physical activity, smoking and alcohol abstinence, weight and stress management, and adherence to a healthy diet were analyzed.

**Results:** A significant threefold increase was observed in the proportion of patients with hypertension who adhered to sodium restriction compared with 20 years ago. However, 70% of patients with hypertension consume more sodium than recommended. Moreover, potassium intake has steadily decreased since 2014, with only 23.8% of patients with hypertension meeting the recommended intake. The body mass index (BMI) and waist circumference of patients with hypertension have gradually increased, with fewer patients maintaining an appropriate weight. The neglect of diet and weight control among young patients with hypertension who experience high stress levels poses challenges in modifying their lifestyles.

**Conclusions:** Patients with hypertension in Korea still consume high amounts of sodium, whereas potassium intake is gradually decreasing. Additionally, obesity rates have been increasing, especially among young patients with hypertension. Considering the increasing number of young patients with hypertension and the low control rate, the importance of lifestyle modifications should be further emphasized.

## Introduction

Hypertension is a leading global health risk and a major independent risk factor for cardiovascular diseases.^1,2^ Globally, more than one-third of adults have hypertension, and its prevalence is increasing owing to global aging.^2,3^ In Korea, the awareness and control of hypertension in recent decades have improved. The awareness and treatment rates for hypertension have increased more than threefold, and the control rate among patients receiving management for hypertension has increased more than tenfold, reaching 70% recently.^4–7^ However, the control rate has stagnated, and the number of hypertension-related deaths has increased by 56.1% over the past decade.^8^ Despite advances in pharmacological treatment, hypertension remains the leading cause of premature death. Therefore, efforts to raise the stagnant control rate of hypertension are required.

Lifestyle modification is a cornerstone of hypertension management independent of medical treatment. Current guidelines recommend lifestyle modification for preventing and treating hypertension.^9–12^ These guidelines recommend effective lifestyle interventions for lowering blood pressure (BP), including regular physical exercise, abstinence from alcohol and smoking, weight management, effective stress management, and healthy dietary modifications with low sodium and high potassium intake. However, lifestyle modifications are often overlooked in hypertension treatment, and data on adherence and changing trends in lifestyle modifications among patients with hypertension do not exist. Therefore, this study aimed to evaluate whether lifestyle modification is effectively implemented and the changes in lifestyle modification trends among patients with hypertension over the past 20 years using data from the Korea National Health and Nutrition Examination Survey (KNHANES) conducted between 1998 and 2021.

## Methods

### Data source

KNHANES is a nationwide representative cross-sectional survey conducted by the Korea Centers for Disease Control and Prevention. This survey has assessed the general health and nutritional statuses of Koreans since 1998.^13^ The first three surveys (1998, 2001, and 2005) were conducted once every 3 years, and it has been conducted annually since 2007. The number of participants for the first three surveys (1998, 2001, and 2005) was approximately 35,000 in each survey. The annual survey included approximately 10,000 participants yearly, except for 2007 when only 5,000 were included. The survey utilizes a complex, stratified, multistage, cluster sampling design. Sample weights are applied to represent the Korean population, considering the complex survey design, survey non-response, and post-stratification. The KNHANES included participants from all age groups; however, our study included individuals aged 20 years and above. The Institutional Review Board of the Chung-Ang University Gwangmyeong Hospital approved the study protocol, which complied with the principles of the Declaration of Helsinki. The requirement for informed consent was waived because of the retrospective nature of the study.

### Variables

The KNHANES included health examinations, health interviews, and nutritional surveys. Trained medical personnel conducted the health examinations using standardized protocols and periodically calibrated all equipment. BP was measured three times on the participant’s right arm using a suitably sized arm cuff and a mercury sphygmomanometer (Baumanometer; WA Baum Co., New York, NY, USA). The measurements were obtained after the participant rested in a seated position for at least 5 min.^12^ The average of the second and third measurements was calculated as the final BP value. Hypertension was defined as systolic BP (SBP) ≥ 140 mmHg, diastolic BP (DBP) ≥ 90 mmHg, or the use of anti-hypertensive drugs.^13,14^ Diabetes mellitus (DM) was defined as fasting blood glucose ≥ 126 mg/dL, glycated hemoglobin (HbA1c) ≥ 6.5%, or on oral hypoglycemic or insulin therapy.^13,14^ Dyslipidemia was defined as total cholesterol level ≥ 240 mg/dL or lipid-lowering medication use. ^13,14^

Trained medical staff conducted the health interviews in-person using standardized questionnaires. Information collected through the interviews included individuals’ age, sex, medical conditions, smoking status, alcohol use, physical activity, mental health, oral health, and weight control. Appropriate body weight control was defined as body mass index (BMI) ≤ 25 kg/m^2^. Moderate alcohol consumption was defined as ≤ 2 drinks and ≤ 1 drink daily for men and women, respectively, following guidelines.^9–12^ Regular physical activity was defined as ≥150 min/week of moderate-intensity aerobic activity, ≥75 min/week of vigorous-intensity aerobic physical activity, or an equivalent combination.

A nutritional survey assessed dietary behaviors, food frequency, and food intake using the 24-h recall method. As recommended by several guidelines, dietary sodium restriction was defined as consuming ≤ 2,000 mg/day, whereas dietary potassium intake was defined as consuming > 3,500 mg/day.^9–11^ The dietary approaches to stop hypertension (DASH) score, developed by Fung et al,^15^ was used to evaluate participants’ adherence to a healthy diet. The DASH score, which evaluates adherence to the DASH diet, is based on the consumption of foods and nutrients emphasized or reduced in the DASH diet.^16^ The DASH score comprises eight components (including fruits, vegetables, nuts and legumes, whole grains, low-fat dairy products, sodium, red and processed meats, and sweetened beverages). The scoring system assigns points ranging from 1 (lowest quintile) to 5 (highest quintile) based on sex quintiles. A score of 1 was assigned to low intake of healthy components (fruits, vegetables, nuts and legumes, whole grains, and low-fat dairy products), whereas the top quintiles receive a score of 5. Conversely, for unhealthy components (including red and processed meat, sugar-sweetened beverage, and sodium intake), the lowest quintile receives 5 points, whereas the highest receives 1 point. The overall score ranges from 8 (lowest adherence) to 40 (highest adherence). As low-fat dairy food intake was not surveyed in the KNHANES, whole dairy intake was used instead.

### Statistical analysis

Sampling weights accounting for the sample design of each KNHANES were used for all statistical analyses. Continuous variables are presented as mean ± standard deviation, whereas categorical variables are reported as relative frequencies (percentages). Statistical analyses were performed using IBM SPSS Statistics v25.0 (IBM Corporation, Armonk, NY, USA) and R programming version 4.0.4 (http://www.R-project.org; R Foundation for Statistical Computing, Vienna, Austria).

## Results

### Baseline Characteristics

Table 1 presents the profile of participants with hypertension during the study. The mean age of the participants with hypertension in 2021 was 61.0 years, revealing a gradual increase from 54.2 years in 1998. Similarly, the mean BMI and waist circumference of those with hypertension gradually increased, from 24.3 kg/m^2^ and 84.9 cm in 1998 to 25.4 kg/m^2^ and 89.3 cm, respectively, in 2021. Furthermore, the participants’ mean SBP and DBP gradually decreased, with mean values of 133.0 mmHg and 80.4 mmHg, respectively. The prevalence of DM and dyslipidemia among participants with hypertension has increased from 19.8% and 20.6% in 2017 to 29.4% and 42.7%, respectively, in 2021.

**Table 1.** Characteristics of subjects with hypertension.

|  | KNHANES I | KNHANES II | KNHANES III | KNHANES IV |  |  | KNHANES V |  |  | KNHANES VI |  |  | KNHANES VII |  |  | KNHANES VIII |  |  |
| --- | --- | --- | --- | --- | --- | --- | --- | --- | --- | --- | --- | --- | --- | --- | --- | --- | --- | --- |
|  | 1998~2000<br>(n=27,201) | 2001~2004<br>(n=26,854) | 2005~2006<br>(n=25,161) | 2007<br>(n=3,304) | 2008<br>(n=7,108) | 2009<br>(n=7,798) | 2010<br>(n=6,664) | 2011<br>(n=6,500) | 2012<br>(n=6,229) | 2013<br>(n=6,028) | 2014<br>(n=5,897) | 2015<br>(n=5,855) | 2016<br>(n=6,315) | 2017<br>(n=6,458) | 2018<br>(n=6,424) | 2019<br>(n=6,542) | 2020<br>(n=6,072) | 2021<br>(n=5,897) |
| Number of subjects | 7,214 | 6,466 | 5,786 | 651 | 1,521 | 1,669 | 1,446 | 1,554 | 1,483 | 1,350 | 1,209 | 1,317 | 1,497 | 1,440 | 1,510 | 1,472 | 1,415 | 1,369 |
| Age, years | 54.2 ± 0.4 | 58.2 ± 0.3 | 58.7 ± 0.3 | 57.9 ± 0.7 | 56.5 ± 0.5 | 55.4 ± 0.5 | 56.1 ± 0.5 | 57.3 ± 0.5 | 57.8 ± 0.6 | 58.0 ± 0.5 | 59.6 ± 0.5 | 59.6 ± 0.5 | 59.2 ± 0.4 | 59.1 ± 0.6 | 59.4 ± 0.5 | 60.3 ± 0.5 | 59.7 ± 0.5 | 61.0 ± 0.5 |
| Male, % | 49.7 | 47.6 | 47.9 | 52.6 | 52.5 | 57.7 | 56.3 | 55.4 | 52.3 | 54.8 | 53.1 | 53.4 | 55.5 | 56.2 | 54.5 | 52.1 | 56.7 | 53.5 |
| Anthropometric measurement |  |  |  |  |  |  |  |  |  |  |  |  |  |  |  |  |  |  |
| Body mass index (kg/m<sup>2</sup>) | 24.3 ± 0.1 | 24.7 ± 0.1 | 25.0 ± 0.1 | 25.4 ± 0.2 | 25.0 ± 0.1 | 25.0 ± 0.1 | 25.0 ± 0.1 | 24.9 ± 0.1 | 25.2 ± 0.1 | 25.4 ± 0.1 | 25.0 ± 0.1 | 25.4 ± 0.1 | 25.4 ± 0.1 | 25.3 ± 0.1 | 25.4 ± 0.1 | 25.3 ± 0.1 | 25.8 ± 0.1 | 25.4 ± 0.1 |
| Waist circumference (cm) | 84.9 ± 0.3 | 86.2 ± 0.3 | 86.2 ± 0.3 | 87.4 ± 0.4 | 86.4 ± 0.3 | 85.6 ± 0.3 | 85.9 ± 0.3 | 86.2 ± 0.4 | 85.8 ± 0.4 | 85.9 ± 0.3 | 85.6 ± 0.3 | 87.5 ± 0.3 | 87.7 ± 0.3 | 86.9 ± 0.3 | 87.5 ± 0.2 | 89.1 ± 0.3 | 90.2 ± 0.3 | 89.3 ± 0.3 |
| SBP (mmHg) | 148.3 ± 0.5 | 146.1 ± 0.6 | 138.5 ± 0.6 | 136.1 ± 0.9 | 133.1 ± 0.6 | 136.7 ± 0.5 | 137.5 ± 0.5 | 135.0 ± 0.5 | 134.8 ± 0.6 | 133.7 ± 0.6 | 132.4 ± 0.6 | 133.9 ± 0.5 | 132.9 ± 0.5 | 132.7 ± 0.5 | 133.2 ± 0.5 | 133.7 ± 0.4 | 133.0 ± 0.5 | 133.0 ± 0.5 |
| DBP (mmHg) | 90.7 ± 0.3 | 89.4 ± 0.4 | 87.4 ± 0.4 | 85.4 ± 0.5 | 85.0 ± 0.4 | 88.2 ± 0.4 | 86.6 ± 0.4 | 83.8 ± 0.4 | 83.4 ± 0.5 | 82.5 ± 0.5 | 81.3 ± 0.5 | 81.4 ± 0.4 | 81.7 ± 0.4 | 81.5 ± 0.5 | 81.9 ± 0.5 | 81.3 ± 0.4 | 81.9 ± 0.4 | 80.4 ± 0.4 |
| Comorbidities |  |  |  |  |  |  |  |  |  |  |  |  |  |  |  |  |  |  |
| Diabetes mellitus, % | - | - |  | 19.8 ± 1.9 | 18.7 ± 1.0 | 17.2 ± 0.9 | 16.8 ± 1.0 | 19.4 ± 1.2 | 18.1 ± 1.2 | 22.1 ± 1.5 | 21.6 ± 1.4 | 19.1 ± 1.2 | 22.3 ± 1.1 | 24.8 ± 1.3 | 23.2 ± 1.1 | 26.1 ± 1.2 | 28.0 ± 1.3 | 29.4 ± 1.2 |
| Dyslipidemia, % | - | - |  | 20.6 ± 1.9 | 18.3 ± 0.1 | 17.3 ± 0.1 | 21.1 ± 1.0 | 22.0 ± 1.3 | 24.0 ± 1.3 | 25.4 ± 1.4 | 25.7 ± 1.5 | 29.7 ± 1.2 | 32.3 ± 1.4 | 37.0 ± 1.4 | 36.2 ± 1.4 | 39.0 ± 1.5 | 41.5 ± 1.4 | 42.7 ± 1.4 |
| Lifestyle behavior |  |  |  |  |  |  |  |  |  |  |  |  |  |  |  |  |  |  |
| Daily calorie intake (kcal) | 1916.5 ± 25.8 | 1849.9 ± 29.9 | 1914.1 ± 26.4 | 1725.6 ± 49.8 | 1797.5 ± 25.8 | 1894.5 ± 25.9 | 2012.5 ± 28.1 | 1988.2 ± 38.2 | 1911.4 ± 32.0 | 1973.2 ± 32.1 | 1968.1 ± 36.0 | 1955.9 ± 31.5 | 1940.4 ± 33.2 | 1889.4 ± 29.4 | 1912.0 ± 30.6 | 1804.3 ± 23.9 | 1846.7 ± 30.0 | 1756.4 ± 29.7 |
| Daily sodium consumption (mg) | 4932.4 ± 102.2 | 5465.5 ± 107.3 | 5516.9 ± 104.1 | 4600.4 ± 171.1 | 4648.8 ± 103.0 | 4885.6 ± 87.3 | 4998.7 ± 86.3 | 4988.5 ± 124.3 | 4595.1 ± 117.2 | 3879.3 ± 98.6 | 3766.8 ± 91.0 | 3650.6 ± 76.0 | 3269.5 ± 82.1 | 3350.0 ± 76.1 | 3281.3 ± 65.4 | 3191.2 ± 56.4 | 3316.2 ± 68.3 | 3154.3 ± 64.7 |
| Daily potassium consumption (mg) | 2606.1 ± 47.5 | 2864.1 ± 55.0 | 2817.4 ± 47.3 | 2678.8 ± 73.5 | 2858.8 ± 50.6 | 2932.2 ± 45.8 | 3050.8 ± 49.7 | 2952.8 ± 59.8 | 2958.6 ± 65.8 | 2935.9 ± 57.7 | 3099.4 ± 57.3 | 2911.7 ± 54.7 | 2875.2 ± 52.9 | 2765.2 ± 44.4 | 2676.3 ± 43.4 | 2688.3 ± 38.2 | 2728.5 ± 46.0 | 2698.6 ± 50.6 |
| Laboratory findings |  |  |  |  |  |  |  |  |  |  |  |  |  |  |  |  |  |  |
| Total cholesterol (mg/dL) | 199.4 ± 1.0 | 199.4 ± 1.0 | 194.0 ± 1.0 | 198.7 ± 2.0 | 196.4 ± 1.2 | 193.7 ± 0.9 | 193.2 ± 1.1 | 194.5 ± 1.1 | 192.6 ± 1.2 | 192.6 ± 1.3 | 188.3 ± 1.3 | 192.9 ± 1.1 | 192.5 ± 1.0 | 189.7 ± 1.2 | 189.2 ± 1.0 | 188.9 ± 1.1 | 185.4 ± 1.2 | 181.2 ± 1.3 |
| HDL cholesterol (mg/dL) | 48.5 ± 0.3 | 44.6 ± 0.3 | 43.0 ± 0.3 | 45.8 ± 0.4 | 46.5 ± 0.4 | 45.7 ± 0.3 | 46.3 ± 0.3 | 48.1 ± 0.4 | 47.7 ± 0.4 | 48.3 ± 0.4 | 48.3 ± 0.4 | 48.6 ± 0.4 | 48.2 ± 0.3 | 48.5 ± 0.3 | 48.0 ± 0.3 | 50.6 ± 0.4 | 48.3 ± 0.4 | 49.0 ± 0.4 |
| LDL cholesterol (mg/dL) | 122.8 ± 0.9 | 121.5 ± 1.0 | 119.1 ± 0.9 | - | - | 116.4 ± 1.6 | 115.9 ± 1.6 | 116.9 ± 1.9 | 118.1 ± 2.5 | 107.8 ± 2.1 | 110.8 ± 2.2 | 114.1 ± 1.0 | 111.4 ± 1.7 | 114.9 ± 2.4 | 112.0 ± 2.1 | 114.3 ± 2.6 | 106.3 ± 2.1 | 107.4 ± 2.4 |
| Triglyceride (mg/dL) | 145.0 ± 1.7 | 167.2 ± 2.6 | 178.3 ± 4.8 | 163.5 ± 4.0 | 173.7 ± 4.3 | 172.3 ± 4.0 | 166.1 ± 3.5 | 166.5 ± 4.1 | 165.2 ± 3.8 | 177.6 ± 5.7 | 165.0 ± 3.7 | 166.9 ± 4.0 | 178.1 ± 6.2 | 160.3 ± 4.2 | 167.2 ± 4.1 | 152.3 ± 3.1 | 164.6 ± 4.9 | 152.2 ± 3.9 |
| Fasting glucose (mg/dL) | 108.1 ± 1.1 | 10.29 ± 0.6 | 102.8 ± 0.7 | 105.0 ± 1.4 | 105.8 ± 0.8 | 104.6 ± 0.7 | 103.6 ± 0.7 | 104.3 ± 0.7 | 105.0 ± 0.9 | 106.5 ± 0.8 | 107.8 ± 0.9 | 107.9 ± 0.9 | 109.7 ± 0.8 | 110.0 ± 0.8 | 109.4 ± 0.7 | 108.8 ± 0.7 | 109.2 ± 0.8 | 111.7 ± 0.9 |
| Creatinine (mg/dL) | 0.93 ± 0.01 | 1.01 ± 0.01 | 1.03 ± 0.01 | 1.01 ± 0.01 | 0.94 ± 0.01 | 0.89 ± 0.01 | 0.87 ± 0.01 | 0.89 ± 0.01 | 0.88 ± 0.01 | 0.94 ± 0.03 | 0.88 ± 0.01 | 0.91 ± 0.02 | 0.91 ± 0.01 | 0.89 ± 0.01 | 0.85 ± 0.01 | 0.87 ± 0.01 | 0.87 ± 0.01 | 0.86 ± 0.01 |

KNHANES, Korea National Health and Nutrition Examination Survey; SBP, systolic blood pressure; DBP, diastolic blood pressure; HDL, high density lipoprotein; LDL, low density lipoprotein

### Dietary Habits

The mean daily sodium intake of participants with hypertension initially increased from 4932.4 mg to 5516.9 mg between 1998 and 2006 before gradually decreasing to 3154.3 mg/day in 2021. Women had lower daily sodium intake than men (2452.8 mg/day for women and 3827.7 mg/day for men in 2021) (Supplementary Tables 1 and 2). A slight increase was observed in the daily potassium intake until 2010 (from 2606.1 mg/day in 1998 to 3050.8 mg/day in 2010), followed by a gradual decrease to 2698.6 mg/day in 2021.

The proportion of participants with hypertension who had adequate sodium restriction as recommended by the guidelines at < 2,000 mg/day was 11.7% in 1998, gradually increasing to 30.4% by 2021 (Fig 1-A). Women have improved in sodium restriction adherence over the decades, increasing from 18.7% in 1998 to 47.1% in 2021. However, men had a lower improvement rate reaching 14.2% in 2021. When stratified into age groups, the proportion of participants meeting the recommended sodium intake increased across all three age groups over the last two decades; however, those in the young (20–39 years) and middle-age (40–59 years) groups were only 18.9% and 25.7% in 2021, respectively (Fig 3-A).

**Figure 1.**
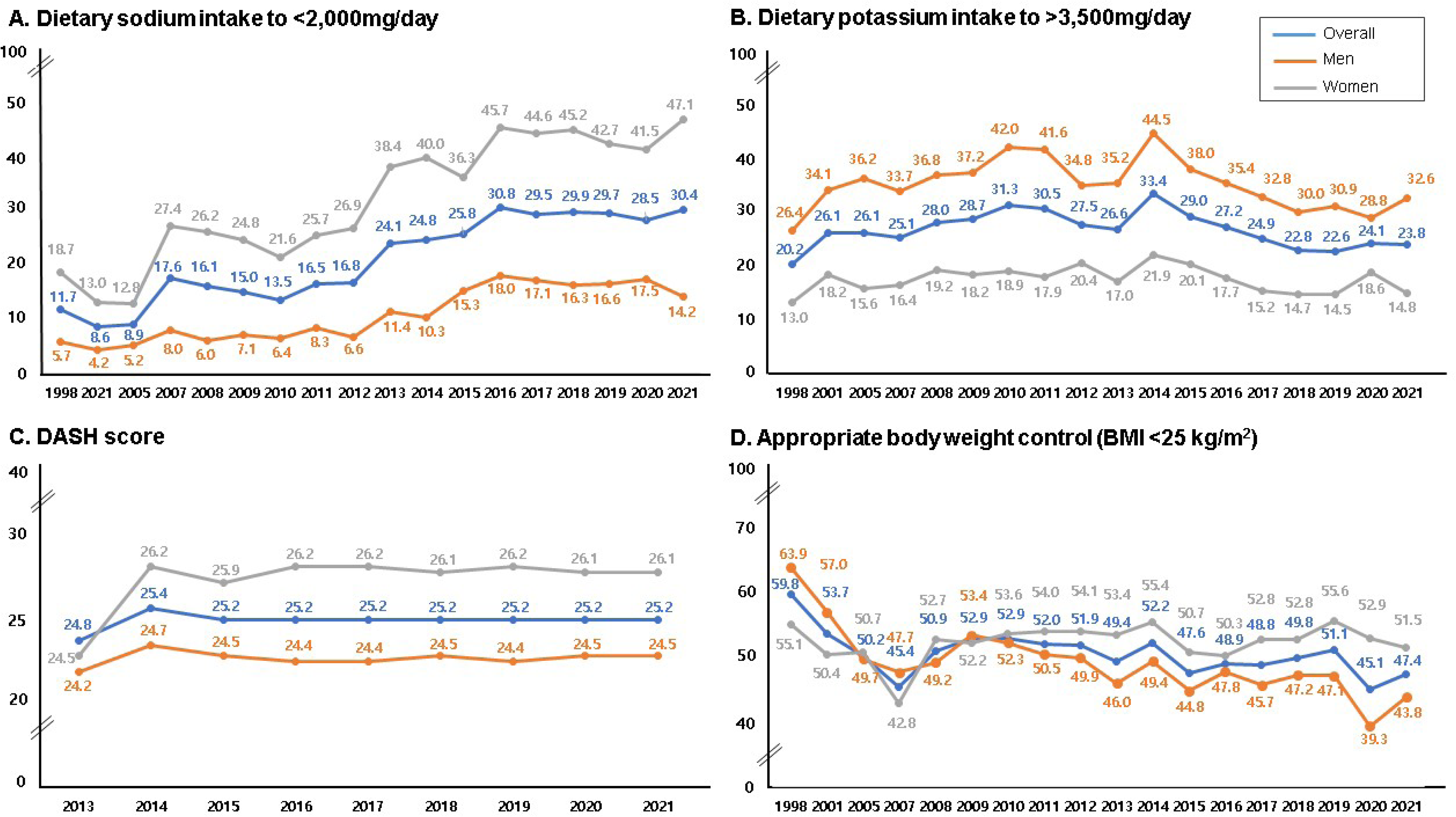
Temporal trends in lifestyle modification in patients with hypertension. (A) Dietary sodium intake <2,000 mg/day. (B) Dietary potassium intake >3,500 mg/day. (C) DASH score. (D) Appropriate body weight control (BMI <25 kg/m^2^). DASH, Dietary Approaches to Stop Hypertension; BMI, body mass index

The percentage of participants with hypertension meeting the recommended potassium intake (>3500 mg/day) increased from 20.2% in 1998 to 33.4% in 2014, but gradually declined to 23.8% in 2021. Men had a higher rate of adequate potassium intake than women (32.6% for men and 14.8% for women in 2021) (Fig 1-B). Among the young participants with hypertension, the proportion that met the recommended potassium intake, which increased to 40% in the 2000s, gradually decreased in the 2010s, becoming 23.8% by 2021. The potassium intake among those aged 60 and older plateaued at approximately 20%, except in 2014 and 2015 (Fig 3-B).

The DASH score plateaued at approximately 25 points from 2013 to 2021 (Fig 1-C). Women had approximately 2 points higher DASH score than men (26.1 and 24.5 points, respectively), with the old-age group having the highest DASH score (26.7, 24.7, and 21.7 points in the old, middle, and young age groups, respectively, in 2021) (Fig 3-C).

### Weight Control

The rate of maintaining an appropriate body weight (BMI <25 kg/m^2^) among patients with hypertension remained at approximately 50% for 20 years (Fig 1-D). More women had appropriate body weights than men. The hypertension prevalence with BMI <25 kg/m^2^ among the older (over 60 years) and middle-aged groups plateaued at approximately 50% and 40%, respectively; however, among the young-age group, the rate of those with BMI <25 kg/m^2^ gradually decreased over the past 20 years, reaching 22.3% by 2021 (Fig 3-D).

### Drinking and Smoking

Overall, more than 60% of the participants with hypertension reported consuming alcohol within the recommended amount in the guidelines.^9–12^ Until the mid-2010s, the abstinence rate among women regarding alcohol consumption exceeded 80%. However, the number of women engaging in heavy drinking gradually increased in the late 2010s. The rate of moderate drinking among men with hypertension increased from 49.6% to 60% between 2012 and 2021 (Fig 2-A). Among the older-age group, more than 70% reported an appropriate amount of drinking. The moderate drinking rate of middle-aged individuals with hypertension steadily increased from 51.9% in 2008 to 61.8% in 2021. The moderation in drinking among young patients with hypertension has recently decreased over the past 2 years, reaching 52.0% in 2021 (Fig 4-A).

**Figure 2.**
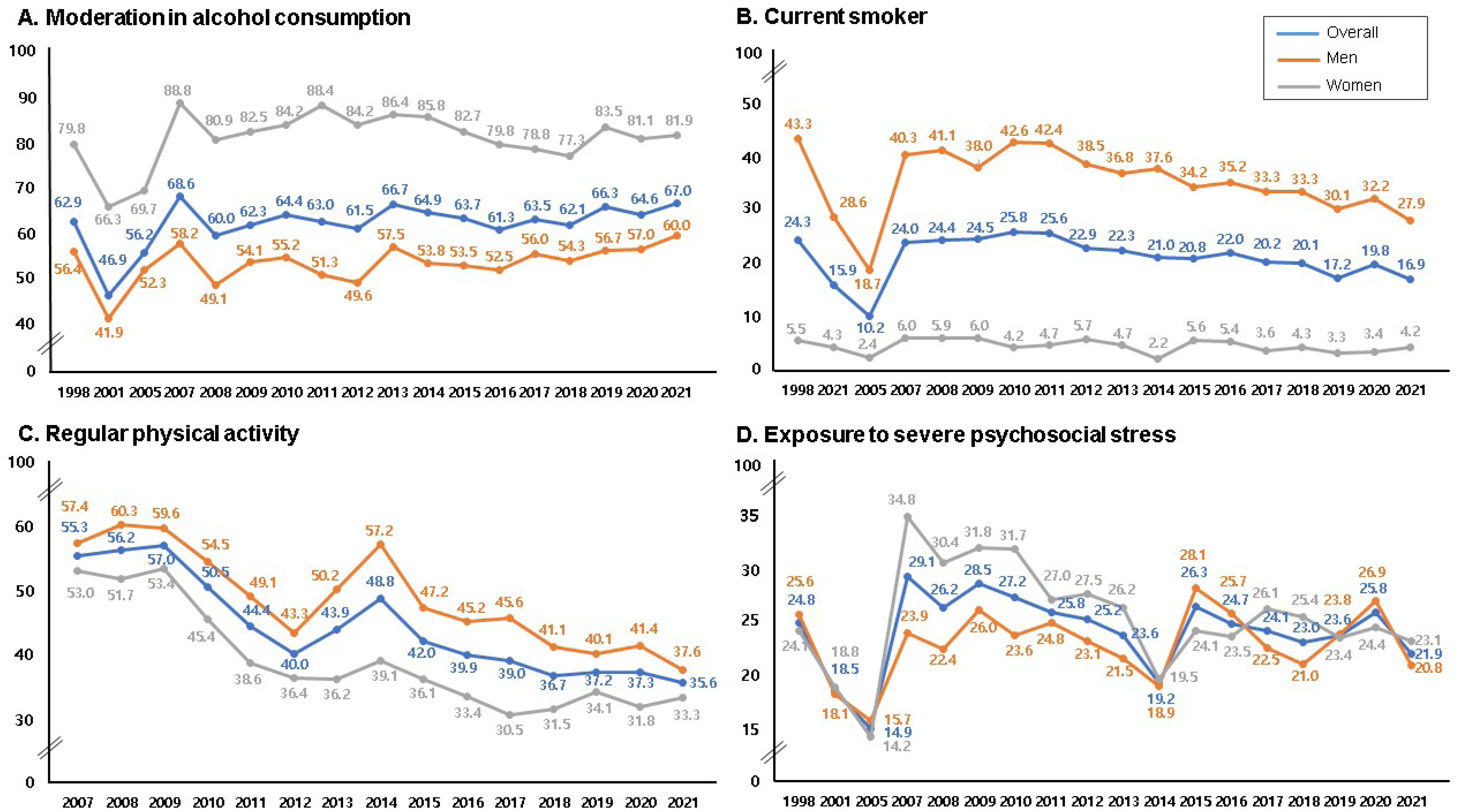
Temporal trends in lifestyle modification in patients with hypertension. (A) Moderation in alcohol consumption. (B) Current smoking status. (C) Regular physical activity. (D) Exposure to severe psychosocial stress.

**Figure 3.**
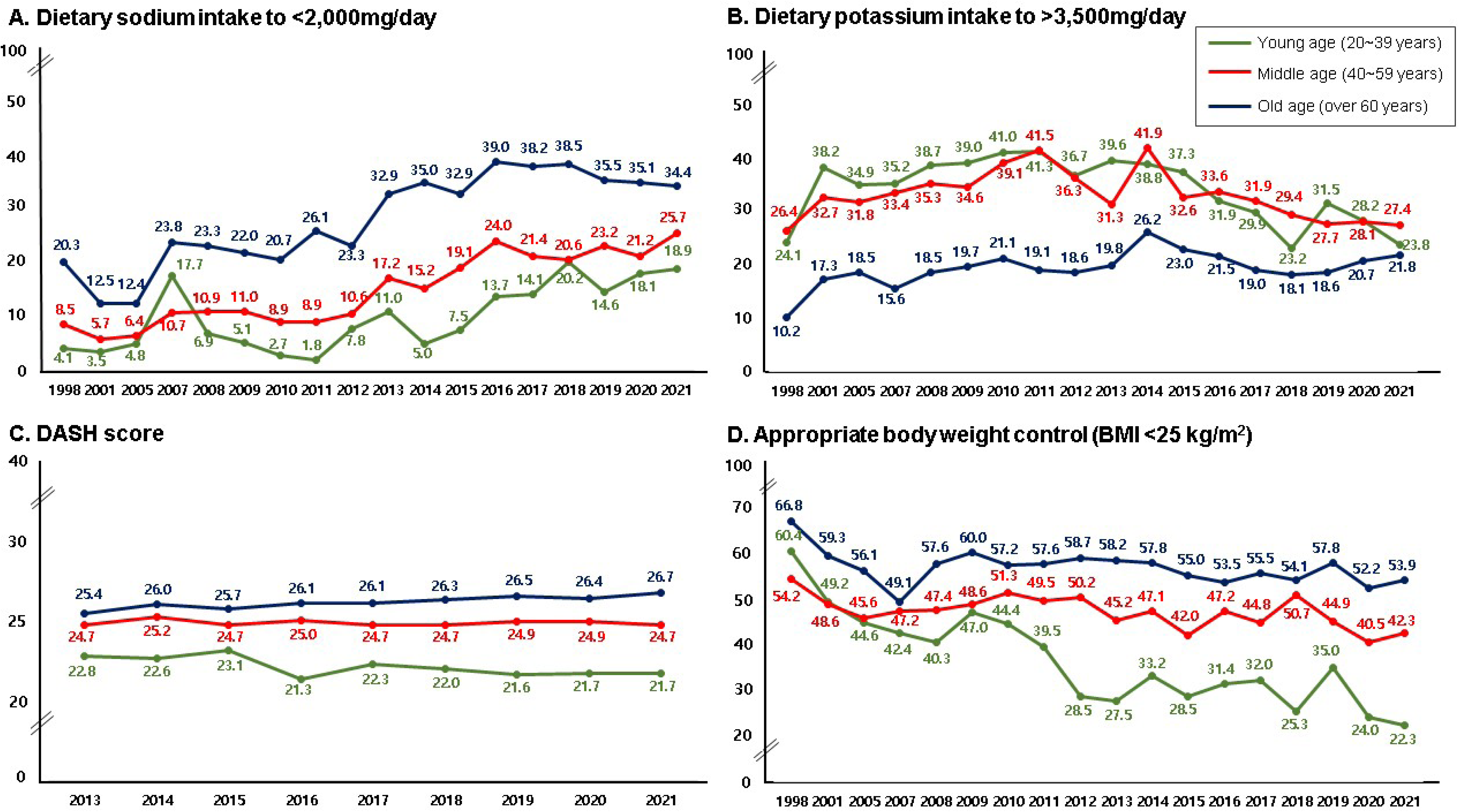
Temporal trends in lifestyle modification in patients with hypertension based on age. (A) Dietary sodium intake<2,000 mg/day. (B) Dietary potassium intake>3,500 mg/day. (C) DASH score. (D) Appropriate body weight control (BMI <25 kg/m^2^). DASH, Dietary Approaches to Stop Hypertension; BMI, body mass index

**Figure 4.**
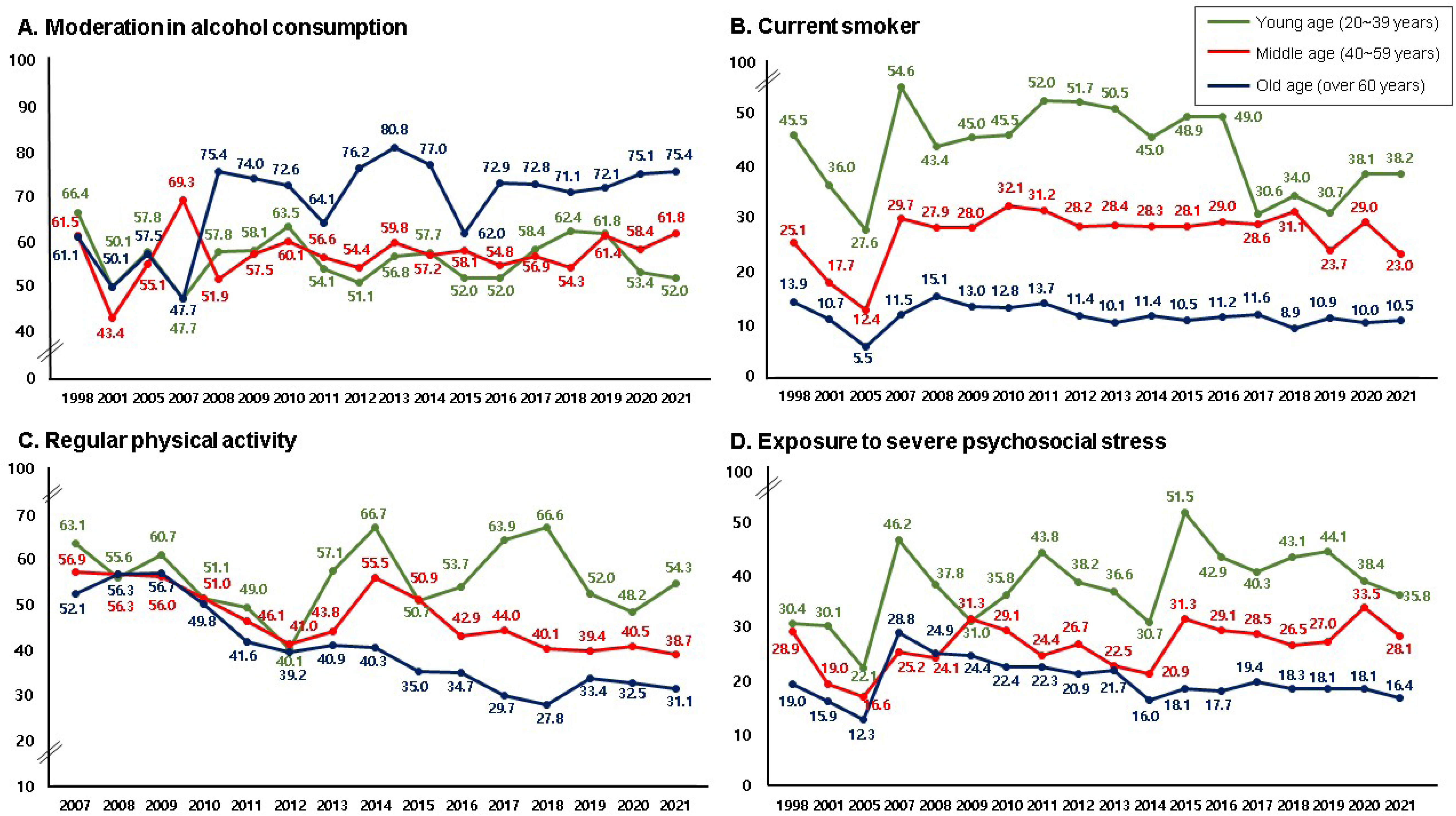
Temporal trends in lifestyle modification in patients with hypertension based on age. (A) Moderation in alcohol consumption. (B) Current smoking status. (C) Regular physical activity. (D) Exposure to severe psychosocial stress.

The rate of current smokers has gradually decreased since 2011, reaching 16.9% by 2021. Smoking rates are relatively low among Korean women, whereas more than a quarter of men are still smokers, although the smoking rate is declining (Fig 2-B). The smoking prevalence among participants with hypertension aged ≥60 years gradually decreased over the 20-year period, reaching 10.5% in 2021. However, the smoking rate among the young participants exceeded 50% from 2011 to 2013 (52.0%, 51.7%, and 50.5%, respectively), remaining at 30% since 2017 (Fig 4-B).

### Regular Physical Activity and Psychosocial Stress

The rate of regular physical activity, which was 57.0% in 2009, decreased annually, except in 2013 and 2014, reaching 35.6% in 2021. Women with hypertension (33.3%) engaged in less regular physical activities than did men (37.6%) in 2021 (Fig 2-C). Regular physical activity among the young-age participants with hypertension decreased to 40.1% in 2012, increased to 66.7% and 66.6% in 2014 and 2018, respectively, and declined in the following 3 years. The rate of regular physical activity decreased annually in middle- and old-age participants. Regular physical activity in middle-aged individuals peaked at 56.9% in 2007 and reached its lowest point at 38.7% in 2021. The rate of regular physical activity rapidly decreased in the old-age group, decreasing from its peak of 56.3% in 2008 to 31.1% by 2021 (Fig 4-C).

In 2021, 21.9% of patients with hypertension (23.1% of men and 20.0% of women) reported experiencing severe psychosocial stress (Fig 2-D). The proportion of young participants with hypertension experiencing severe stress declined from 51.5% in 2015 to 35.8% in 2021. In contrast, the proportion of participants experiencing severe stress remained relatively constant over 20 years among middle-aged (28.1%) and old-age (16.4%) participants in 2021 (Fig 4-D).

## Discussion

Despite the increasing emphasis on the importance of lifestyle modifications for effectively managing hypertension, this study demonstrated that lifestyle modifications are still poorly implemented. This study revealed the following: 1) The proportion of patients with hypertension adhering to sodium restriction in 2021 increased significantly threefold compared with 20 years ago; however, 7 out of 10 patients with hypertension still consumed more sodium than the recommended amount. Potassium intake has decreased since 2014, with only 22.8% of patients with hypertension meeting the recommended intake; 2) the BMI and waist circumference of patients with hypertension have gradually increased, whereas the rate of patients with hypertension maintaining an appropriate weight has gradually decreased; 3) regular physical activity in middle-aged and old-age has decreased; 4) young patients with hypertension have neglected diet and weight control and experienced severe stress, making it challenging to modify their lifestyle.

This study is the first to demonstrate the trends in lifestyle modifications in patients with hypertension from 1998 to 2018 in Korea. Therefore, these findings will provide valuable insights into the current status of lifestyle modifications in patients with hypertension and determine further strategies for promoting lifestyle interventions.

### Prevalence, awareness, and treatment of hypertension in Korea

The prevalence of hypertension in Korea remained stable at 29% from 1998 to 2021. However, the awareness and treatment of hypertension improved during the study period. The awareness and treatment rates of hypertension increased over three-fold, and the control rates increased over ten-fold between 1998 and 2021.^5,17^ However, despite many advances, the control rate of hypertension in 2021 was only 47.4% and has stagnated over the past decade.^17^ This highlights the need for continued attention and practice of lifestyle modification, in addition to medical treatment, to increase the control rates of hypertension.

### Sodium Restriction and Increased Potassium Intake

Sodium restriction and increased potassium intake are the most fundamental recommendations for dietary interventions in hypertension. Meta-analyses have revealed that these interventions effectively reduce BP,^18,19^ with a linear dose-response relationship observed.^19,20^ The relationship between sodium restriction, increased potassium intake, and BP is stronger in individuals with SBP ≥ 140 mmHg.^21,22^ Potassium suppresses BP elevation caused by excessive salt intake by promoting sodium excretion from the body. The higher the salt intake, the more pronounced the BP-lowering effect of potassium. These findings suggest that more aggressive sodium restriction and potassium intake are required for patients with poorly controlled BP. However, caution should be exercised regarding potassium intake in patients with chronic kidney disease.

Considering the importance and effects of dietary interventions for hypertension, the trends in sodium and potassium intakes among participants with hypertension in our study have implications. Traditionally, Korean food has been associated with higher salt content than that of other countries. However, compared with two decades ago, the overall sodium intake of patients with hypertension has decreased. The proportion of individuals consuming the recommended amount (<2000 mg/day) has tripled. Nevertheless, approximately 70% of individuals with hypertension still exceed the recommended sodium intake. In addition, potassium intake has decreased since the mid-2010s, with only 23.8% of individuals with hypertension meeting the recommended amount by 2021. Overall, 20% of patients with hypertension aged <60 years adhere to adequate salt restriction. Therefore, continuous attention and education are necessary to improve dietary habits.

### Appropriate Weight Management and Enhanced Physical Exercise

Furthermore, we observed that the BMI and waist circumference of individuals with hypertension have gradually increased over the past 20 years. Moreover, the proportion of patients maintaining an appropriate BMI has gradually decreased. The increase in BMI was particularly pronounced in young patients with hypertension. Additionally, physical exercise has decreased among patients with hypertension. The rate of regular physical exercise, which was 55.3% in 2007, gradually decreased to 35.6% by 2021. The decrease in regular physical activity was remarkable in patients with hypertension aged >40 years. This suggests the need for an energy-restricted diet in young patients and regular exercise in older patients.

A meta-analysis of 25 RCTs revealed that weight reduction lowers BP. The analysis demonstrated that SBP and DBP decreased by approximately 1 mmHg for each 1 kg body weight loss, with or without hypertension.^23^ Weight loss is accompanied by a decrease in visceral fat (a risk factor for hypertension), a decrease in renin-angiotensin-aldosterone system activity, improved vascular endothelial function, and reduced systemic inflammation.^24^

Proper dietary restrictions are important for weight loss; however, exercise should be combined for added benefits. One study demonstrated greater BP reduction when exercise was combined with a diet and weight management program (SBP of 7.4 mmHg and DBP of 5.6 mmHg, respectively). In addition, BP decreased considerably even in the group with minimal weight loss after exercise intervention.^25^ Therefore, physical exercise benefits weight loss and BP control in patients with hypertension. Encouraging continuous exercise and developing accessible exercise programs for the elderly are essential to promote physical activity.

### Lifestyle Modification for Young Patients with Hypertension

The young participants with hypertension exhibited low DASH scores, high salt intake, insufficient potassium intake, high BMI, high smoking rates, and high rates of severe psychosocial stress. These findings indicate a growing trend towards modernized lifestyles among young patients with hypertension. A modernized lifestyle characterized by sedentary behavior, high intake of processed meat and alcohol, imbalanced sodium-to-potassium ratio, and chronic psychosocial stress may be associated with an increased hypertension prevalence and lower control rates among young patients with hypertension, leading to an increased risk of cardiovascular diseases in the future. Therefore, improving awareness and education about lifestyle modifications among young patients with hypertension is necessary.

### Further Prospectives of Improvement in Lifestyle Modification

Increasing the awareness of the importance of lifestyle modification and drug treatment for long-term BP control in patients with hypertension and prevention of cardiovascular disease is necessary for proper lifestyle optimization. In addition, clinicians should provide personalized guidelines to encourage lifestyle changes and motivate patients with hypertension to adopt lifestyle changes. Further research is required to explore the factors and methods that can facilitate the maintenance of a healthy lifestyle. A recent systematic review and meta-analysis revealed that application-based interventions can aid patients in making healthy nutrition behavior changes and improving nutrition-related health outcomes.^26,27^ Based on the HERB-DH1 pivotal study, feedback through an application enhanced the effectiveness of lifestyle modification and lowered the 24-h ambulatory, home, and office BP compared with the standard lifestyle adjustment recommendations.^28^ In the digital therapeutics group using the application, substantial reductions in salt intake and weight were observed, highlighting the effectiveness of the application-based feedback in promoting diet and weight control. Lifestyle modification using the application is beneficial for young patients with hypertension having poor diet and weight control owing to modernized lifestyles. Furthermore, although there are small studies or ongoing RCTs, several studies that have helped lower BP through lifestyle modification using motivational education programs, telehealth, text-message-based lifestyle interventions, and wearable devices should be considered. ^29–31^

### Limitation

This study had some limitations. First, the longitudinal follow-up was limited because the KNHANES is a cross-sectional study. However, KNHANES provides nationally representative data with a complex survey design suitable for understanding the prevalence and trends of hypertension. Second, the questionnaire was modified slightly from KNHANES I to VI; thus, variations in the definitions of each variable were observed during the study period. Lastly, self-reported questionnaires on smoking, alcohol consumption, physical activity, and mental health were used owing to the nature of KNHANES. Differences between reported behavior and reality may have existed because participants may have experienced recall bias or deliberately underestimated or overestimated their actual practice.

### Perspectives

This study reported the trends in lifestyle modifications among patients with hypertension. Enhancing lifestyle habits, including dietary habits, weight management, and increased physical exercise time, is necessary to improve the stagnant control rate of hypertension. In addition, young patients with hypertension are adopting a more modernized lifestyle than they did 20 years ago. Considering the increasing prevalence and low control rate among young patients with hypertension, continuous attention and efforts toward lifestyle modification are required.

### Novelty and Relevance What is new?

Previous studies have highlighted the importance of lifestyle modifications for hypertension management; however, data on the adherence to these modifications and their changing patterns over time in this specific population is lacking. This study is novel because it focuses on evaluating the implementation and trends in lifestyle modifications among patients with hypertension in Korea over a period of two decades.

### What is Relevant?

1. Only 20∼30% of patients with hypertension intake adequate sodium and potassium.
2. The BMI and waist circumference of patients with hypertension have gradually increased and regular physical activity has decreased.
3. Young patients with hypertension have neglected diet and weight control and experienced severe stress, making it challenging to modify their lifestyle.

### Clinical/Pathophysiological Implications?

This study is the first to demonstrate the trends in lifestyle modifications in patients with hypertension. these findings will provide valuable insights into the current status of lifestyle modifications in patients with hypertension and determine further strategies for promoting lifestyle interventions.

## Data Availability

Data from the Korea National Health and Nutrition Examination Survey (KNHANES). KNHANES is a nationwide representative cross-sectional survey conducted by the Korea Centers for Disease Control and Prevention.

## ABBREVIATIONS AND ACRONYMS

KNHANES: Korea National Health and Nutrition Examination Survey
BMI: body mass index
BP: blood pressure
SBP: systolic blood pressure
DBP: diastolic blood pressure
DM: diabetes mellitus
HbA1c: glycated hemoglobin
DASH: Dietary Approaches to Stop Hypertension

## Acknowledgments

none

## References

1. Collaborators GBDRF. Global, regional, and national comparative risk assessment of 84 behavioural, environmental and occupational, and metabolic risks or clusters of risks for 195 countries and territories, 1990-2017: a systematic analysis for the Global Burden of Disease Study 2017. Lancet. 2018;392:1923–1994. doi: 10.1016/S0140-6736(18)32225-6

2. Benjamin EJ, Virani SS, Callaway CW, Chamberlain AM, Chang AR, Cheng S, Chiuve SE, Cushman M, Delling FN, Deo R, et al. Heart Disease and Stroke Statistics-2018 Update: A Report From the American Heart Association. Circulation. 2018;137:e67–e492. doi: 10.1161/CIR.0000000000000558

3. Forouzanfar MH, Liu P, Roth GA, Ng M, Biryukov S, Marczak L, Alexander L, Estep K, Hassen Abate K, Akinyemiju TF, et al. Global Burden of Hypertension and Systolic Blood Pressure of at Least 110 to 115 mm Hg, 1990-2015. JAMA. 2017;317:165–182. doi: 10.1001/jama.2016.19043

4. Guo F, He D, Zhang W, Walton RG. Trends in prevalence, awareness, management, and control of hypertension among United States adults, 1999 to 2010. J Am Coll Cardiol. 2012;60:599–606. doi: 10.1016/j.jacc.2012.04.026

5. Kang SH, Kim SH, Cho JH, Yoon CH, Hwang SS, Lee HY, Youn TJ, Chae IH, Kim CH. Prevalence, Awareness, Treatment, and Control of Hypertension in Korea. Sci Rep. 2019;9:10970. doi: 10.1038/s41598-019-46965-4

6. Neuhauser HK, Adler C, Rosario AS, Diederichs C, Ellert U. Hypertension prevalence, awareness, treatment and control in Germany 1998 and 2008-11. J Hum Hypertens. 2015;29:247–253. doi: 10.1038/jhh.2014.82

7. Roy A, Praveen PA, Amarchand R, Ramakrishnan L, Gupta R, Kondal D, Singh K, Sharma M, Shukla DK, Tandon N, et al. Changes in hypertension prevalence, awareness, treatment and control rates over 20 years in National Capital Region of India: results from a repeat cross-sectional study. BMJ Open. 2017;7:e015639. doi: 10.1136/bmjopen-2016-015639

8. Virani SS, Alonso A, Benjamin EJ, Bittencourt MS, Callaway CW, Carson AP, Chamberlain AM, Chang AR, Cheng S, Delling FN, et al. Heart Disease and Stroke Statistics-2020 Update: A Report From the American Heart Association. Circulation. 2020;141:e139–e596. doi: 10.1161/CIR.0000000000000757

9. Whelton PK, Carey RM, Aronow WS, Casey DE, Jr., Collins KJ, Dennison Himmelfarb C, DePalma SM, Gidding S, Jamerson KA, Jones DW, et al. 2017 ACC/AHA/AAPA/ABC/ACPM/AGS/APhA/ASH/ASPC/NMA/PCNA Guideline for the Prevention, Detection, Evaluation, and Management of High Blood Pressure in Adults: A Report of the American College of Cardiology/American Heart Association Task Force on Clinical Practice Guidelines. J Am Coll Cardiol. 2018;71:e127–e248. doi: 10.1016/j.jacc.2017.11.006

10. Williams B, Mancia G, Spiering W, Agabiti Rosei E, Azizi M, Burnier M, Clement DL, Coca A, de Simone G, Dominiczak A, et al. 2018 ESC/ESH Guidelines for the management of arterial hypertension. Eur Heart J. 2018;39:3021–3104. doi: 10.1093/eurheartj/ehy339

11. Unger T, Borghi C, Charchar F, Khan NA, Poulter NR, Prabhakaran D, Ramirez A, Schlaich M, Stergiou GS, Tomaszewski M, et al. 2020 International Society of Hypertension Global Hypertension Practice Guidelines. Hypertension. 2020;75:1334–1357. doi: 10.1161/HYPERTENSIONAHA.120.15026

12. Lee HY, Shin J, Kim GH, Park S, Ihm SH, Kim HC, Kim KI, Kim JH, Lee JH, Park JM, et al. 2018 Korean Society of Hypertension Guidelines for the management of hypertension: part II-diagnosis and treatment of hypertension. Clin Hypertens. 2019;25:20. doi: 10.1186/s40885-019-0124-x

13. Kweon S, Kim Y, Jang MJ, Kim Y, Kim K, Choi S, Chun C, Khang YH, Oh K. Data resource profile: the Korea National Health and Nutrition Examination Survey (KNHANES). Int J Epidemiol. 2014;43:69–77. doi: 10.1093/ije/dyt228

14. Lee HH, Cho SMJ, Lee H, Baek J, Bae JH, Chung WJ, Kim HC. Korea Heart Disease Fact Sheet 2020: Analysis of Nationwide Data. Korean Circ J. 2021;51:495–503. doi: 10.4070/kcj.2021.0097

15. Fung TT, Chiuve SE, McCullough ML, Rexrode KM, Logroscino G, Hu FB. Adherence to a DASH-style diet and risk of coronary heart disease and stroke in women. Arch Intern Med. 2008;168:713–720. doi: 10.1001/archinte.168.7.713

16. Soltani S, Arablou T, Jayedi A, Salehi-Abargouei A. Adherence to the dietary approaches to stop hypertension (DASH) diet in relation to all-cause and cause-specific mortality: a systematic review and dose-response meta-analysis of prospective cohort studies. Nutr J. 2020;19:37. doi: 10.1186/s12937-020-00554-8

17. Kim HC, Lee H, Lee HH, Seo E, Kim E, Han J, Kwon JY, Korean Society of Hypertension -Hypertension Epidemiology Research Working G. Korea hypertension fact sheet 2021: analysis of nationwide population-based data with special focus on hypertension in women. Clin Hypertens. 2022;28:1. doi: 10.1186/s40885-021-00188-w

18. Huang L, Trieu K, Yoshimura S, Neal B, Woodward M, Campbell NRC, Li Q, Lackland DT, Leung AA, Anderson CAM, et al. Effect of dose and duration of reduction in dietary sodium on blood pressure levels: systematic review and meta-analysis of randomised trials. BMJ. 2020;368:m315. doi: 10.1136/bmj.m315

19. Binia A, Jaeger J, Hu Y, Singh A, Zimmermann D. Daily potassium intake and sodium-to-potassium ratio in the reduction of blood pressure: a meta-analysis of randomized controlled trials. J Hypertens. 2015;33:1509–1520. doi: 10.1097/HJH.0000000000000611

20. Graudal N, Hubeck-Graudal T, Jurgens G, Taylor RS. Dose-response relation between dietary sodium and blood pressure: a meta-regression analysis of 133 randomized controlled trials. Am J Clin Nutr. 2019;109:1273–1278. doi: 10.1093/ajcn/nqy384

21. Graudal NA, Hubeck-Graudal T, Jurgens G. Effects of low sodium diet versus high sodium diet on blood pressure, renin, aldosterone, catecholamines, cholesterol, and triglyceride. Cochrane Database Syst Rev. 2017;4:CD004022. doi: 10.1002/14651858.CD004022.pub4

22. Aburto NJ, Hanson S, Gutierrez H, Hooper L, Elliott P, Cappuccio FP. Effect of increased potassium intake on cardiovascular risk factors and disease: systematic review and meta-analyses. BMJ. 2013;346:f1378. doi: 10.1136/bmj.f1378

23. Neter JE, Stam BE, Kok FJ, Grobbee DE, Geleijnse JM. Influence of weight reduction on blood pressure: a meta-analysis of randomized controlled trials. Hypertension. 2003;42:878–884. doi: 10.1161/01.HYP.0000094221.86888.AE

24. Valenzuela PL, Carrera-Bastos P, Galvez BG, Ruiz-Hurtado G, Ordovas JM, Ruilope LM, Lucia A. Lifestyle interventions for the prevention and treatment of hypertension. Nat Rev Cardiol. 2021;18:251–275. doi: 10.1038/s41569-020-00437-9

25. Blumenthal JA, Sherwood A, Gullette EC, Babyak M, Waugh R, Georgiades A, Craighead LW, Tweedy D, Feinglos M, Appelbaum M, et al. Exercise and weight loss reduce blood pressure in men and women with mild hypertension: effects on cardiovascular, metabolic, and hemodynamic functioning. Arch Intern Med. 2000;160:1947–1958. doi: 10.1001/archinte.160.13.1947

26. Villinger K, Wahl DR, Boeing H, Schupp HT, Renner B. The effectiveness of app-based mobile interventions on nutrition behaviours and nutrition-related health outcomes: A systematic review and meta-analysis. Obes Rev. 2019;20:1465–1484. doi: 10.1111/obr.12903

27. Thomas JG, Bond DS, Raynor HA, Papandonatos GD, Wing RR. Comparison of Smartphone-Based Behavioral Obesity Treatment With Gold Standard Group Treatment and Control: A Randomized Trial. Obesity (Silver Spring). 2019;27:572–580. doi: 10.1002/oby.22410

28. Kario K, Nomura A, Harada N, Okura A, Nakagawa K, Tanigawa T, Hida E. Efficacy of a digital therapeutics system in the management of essential hypertension: the HERB-DH1 pivotal trial. Eur Heart J. 2021. doi: 10.1093/eurheartj/ehab559

29. Islam FMA, Lambert EA, Islam SMS, Islam MA, Biswas D, McDonald R, Maddison R, Thompson B, Lambert GW. Lowering blood pressure by changing lifestyle through a motivational education program: a cluster randomized controlled trial study protocol. Trials. 2021;22:438. doi: 10.1186/s13063-021-05379-2

30. Taher M, Yule C, Bonaparte H, Kwiecien S, Collins C, Naylor A, Juraschek SP, Bailey-Davis L, Chang AR. Telehealth versus self-directed lifestyle intervention to promote healthy blood pressure: a protocol for a randomised controlled trial. BMJ Open. 2021;11:e044292. doi: 10.1136/bmjopen-2020-044292

31. Bolmsjo BB, Wolff M, Nymberg VM, Sandberg M, Midlov P, Calling S. Text message-based lifestyle intervention in primary care patients with hypertension: a randomized controlled pilot trial. Scand J Prim Health Care. 2020;38:300–307. doi: 10.1080/02813432.2020.1794392

